# Older adults’ beliefs about anxiety: A multicultural qualitative study informed by Leventhal’s Common-Sense Model of Self-Regulation

**DOI:** 10.64898/2026.03.13.26348137

**Authors:** Rasha Alkholy, Karina Lovell, Rebecca Pedley, Penny Bee

## Abstract

**Aim:** Anxiety disorders in older adults are commonly underdiagnosed and undertreated, especially among minority ethnic groups. This UK multicultural qualitative study aimed to explore and compare beliefs about anxiety among self-reporting White British, South Asian, African and Caribbean older adults, using Leventhal’s Common-Sense Model.

**Methods:** Individual interviews were conducted with 52 older adults who self-reported anxiety (current or past). Data were managed and analysed using the Framework Method. Professional interpreters facilitated interviews with non-English speakers.

**Findings:** The study incorporated the perspectives of 27 older adults with distressing anxiety and 25 with non-distressing anxiety. Participants’ beliefs mapped onto the illness-related dimensions in Leventhal’s Common-Sense Model. Beliefs about anxiety differed across and within cultural groups, with notable distinctions between participants with distressing and non-distressing anxiety. Those with distressing anxiety neither normalised anxiety nor considered it as an illness trajectory. Overall, participants had a fragmented understanding of anxiety disorders. Specific aspects of older adults’ beliefs were influenced by their salient identities rather than their cultural background. Two new dimensions were identified: aggravating factors, believed to trigger or exacerbate anxiety symptoms; protective factors, believed to alleviate or prevent mental health problems.

**Conclusions:** Applying Leventhal’s Common-Sense Model to anxiety has yielded new insights with significant implications for understanding potential causes of low mental health services use among older adults. Grouping people into broad categories of cultural groups disregards the diversity among individuals within the same group. Cross-cultural research should embrace this diversity and employ nuanced approaches to provide meaningful, person-centred insights into people’s perceptions of illness.

## INTRODUCTION

Anxiety disorders are common in older adults (1), with prevalence rates varying by country sampled, population studied, timeframe considered and assessment method applied (2). These disorders are associated with poor health outcomes, comorbidities (3) and higher healthcare costs (4). Despite their prevalence and negative consequences, late-life anxiety disorders are usually underdiagnosed and untreated; over 70% of adults over 55 in the USA with past-year anxiety disorders do not seek mental health services (5), a figure that markedly increases with advancing age (6). Moreover, other factors contribute to the heightened risk of mental health services underutilisation in this population, among which ethnicity stands out (7).

Suggestions of ethnic disparities in mental healthcare warrant critical investigation if we are to achieve health equity. Researchers have long maintained that culture significantly impacts the perception and help-seeking behaviours for mental health issues (8). Yet, surprisingly, cross-cultural research on anxiety in older adults is still in its infancy, with a notable scarcity in qualitative research.

One theoretical framework that can help us understand how people perceive and manage illness is Leventhal’s Common-Sense Model of Self-Regulation (9). This model posits that, when faced with a health threat, individuals generate cognitive and emotional representations of that threat. Cognitive representations include beliefs about illness identity, cause, timeline, consequences, whether it could be cured or controlled and the degree to which it makes sense to the individual. Emotional representations involve emotional responses to illness, such as fear, anxiety or depression (10). These representations (or illness beliefs) guide the selection, performance and appraisal of coping actions. They are shaped by personal illness experiences, social and cultural knowledge, and observation of others (11).

To understand the reasons behind low mental health services use among older adults with anxiety, especially those from minority ethnic backgrounds, we need to unpack the illness beliefs that inform their help-seeking behaviours. This insight may help us develop more acceptable services. To our knowledge, no study has yet explored older adults’ beliefs about anxiety by applying Leventhal’s Common-Sense Model (9).

## METHODS

### Study design

This multicultural qualitative study aimed to explore and compare beliefs about anxiety among self-reporting White British, South Asian, African and Caribbean older adults in the UK, informed by Leventhal’s Common-Sense Model. We used the generic qualitative approach described by Ritchie and Spencer (12), which relies on a subtle realist ontological position while embracing aspects of interpretivism and pragmatism.

We collaborated with eight public contributors: five White British, two South Asian and one Caribbean, all living in the UK. Seven were 65 years or older, and five had lived experience of common mental health problems. Together we co-created the interview topic guide, recruitment strategies and materials, and an arts-based workshop to enhance recruitment. The public contributors also contributed to data interpretation, provided input on the final thematic framework and co-authored a commentary on their involvement and study findings **(Appendix-1)**.

### Ethical approval

The study received ethical approval from the University of Manchester Research Ethics Committee (Ref: 2021-10461-17766; Date: 05/02/2021).

### Sampling

Participants were eligible if they were 65 years or over; self-identified as “worriers”, had experienced or were experiencing anxiety or “stress”; self-identified as White British, South Asian (Indian, Pakistani, Bangladeshi), African or Caribbean; and lived independently in the UK. Exclusion criteria included those receiving mental health crisis management. We used purposive sampling to ensure adequate representation across the cultural groups and snowball sampling to expand the reach to potentially eligible participants.

### Recruitment strategy

Data collection primarily occurred during the COVID-19 pandemic national lockdowns. We collaborated with relevant gatekeepers, research networks and our public contributors to circulate the study details to potential participants. We posted study adverts on social media platforms (X/Twitter, Facebook), newsletters and websites of the University of Manchester and various organisations. Physical flyers were distributed in local venues (e.g., churches, mosques, pharmacies) to reach digitally excluded older adults. Our public contributors and gatekeepers assisted in identifying and approaching eligible participants.

### Recruitment procedures

To enhance research inclusivity while preserving participant eligibility, we used different terms to describe anxiety in the study advert, such as “stress” and “worrier” and their synonyms in Urdu, Hindi and Bengali. We confirmed eligibility by asking potential participants if they had ever been diagnosed with an anxiety disorder (current or in the past). Those with a clinical diagnosis were eligible to participate.

Those without a diagnosis were informed that this study aimed to understand people’s experiences of ‘anxiety’, ‘stress’ or ‘worry’ that persisted over long periods and impacted daily functioning or caused significant distress; anxiety could revolve around specific issues or be more generalised, including several issues. If participants believed this description applied to them, either currently or in the past, they were invited to participate.

Interested eligible participants were sent the Participant Information Sheet, consent form and three questionnaires: a sociodemographic data form to collect basic sociodemographic data, the Geriatric Anxiety Scale (13) and Geriatric Depression Scale-Short Form (14) to screen for current symptoms of anxiety and depression, respectively. We did not use the total Geriatric Anxiety Scale score to determine eligibility; it was used as a secondary measure to evaluate the effectiveness of our inclusive recruitment strategy in identifying eligible participants.

### Recruitment material

The study advert, Participant Information Sheet, the Geriatric Anxiety Scale (13) and the consent form were professionally translated into Urdu, Hindi and Bengali (proofread and verified for accuracy by an independent professional translator). A native Urdu-speaking public contributor reviewed the Urdu study materials for linguistic appropriateness and acceptability. The English (14), Urdu (15), Hindi (16) and Bengali (17) versions of the Geriatric Depression Scale-Short Form are accessible in the respective open-access articles and the public domain (https://web.stanford.edu/∼yesavage/GDS.html).

### Data collection procedures

Participants completed two interviews: a brief initial interview to audio-record their verbal consent and responses to the three questionnaires, and a semi-structured interview lasting 38 to 160 minutes. Interviews were conducted via phone (n=27), virtually (Zoom; n=13), or in person (n=12: 7 at participants’ homes, 3 in community organisations, 2 in university facilities). Professional interpreters facilitated interviews with non-English speakers.

Semi-structured interviews followed a topic guide **(Appendix-2)** based on Leventhal’s Common-Sense Model (9). It included five open-ended questions with probes to explore participants’ understanding of anxiety, self-help strategies, help-seeking behaviours, barriers and enablers to mental health services use, and outcomes of interest used to appraise their strategies. The topic guide was iteratively adapted to incorporate new topics raised by participants. Participants received a £30 gift voucher for their time.

Interviews were audio recorded using encrypted devices, transcribed verbatim in English and anonymised. Transcripts were verified against recordings for accuracy. Data collection occurred in the UK over 13 months, from April 2021 to May 2022.

### Data analysis

Interview data were managed and analysed using the Framework Method (18), a comparative form of thematic analysis that employs an organising framework of deductively and inductively derived themes for data analysis (19). This method’s highly structured matrix output organises data into columns (subthemes), rows (cases or participants) and cells of ‘summarised data’, offering an overview of the whole dataset while retaining the integrity of each participant’s account, thus facilitating data analysis both by code and case (20). This feature was pertinent to our study since we wanted to compare participants’ perspectives across and within the cultural groups. We used Leventhal’s Common-Sense Model (9) as the a priori organising framework for initial coding, with inductive analysis identifying new dimensions not recognised by the model.

Data analysis was conducted by RA and cross-checked by KL, PB and RP to ensure analytical rigour and coding consistency; discrepancies were resolved by consensus. Our public contributors reviewed carefully selected extracts of anonymised quotes, provided feedback, and verified data interpretation; discrepancies were resolved to ensure consensus among the wider group. Feedback from our public contributors was used to refine the final thematic framework. Data management was supported by NVivo 12 Plus.

Recruitment ceased when data saturation was reached; a stage at which the research team concluded that codes, themes, linkages and interpretations were sufficiently developed, conceptually dense and coherent to meet the study’s objectives. Additional data did not refine deductively identified themes and codes, nor did they identify new themes, codes or linkages. Data saturation was reached after interviewing 14 White British, 16 South Asian and 15 African or Caribbean participants. One to two additional interviews with the African/Caribbean and White British groups, respectively, confirmed this judgement. Four additional interviews were conducted with the South Asian group to incorporate a more linguistically diverse sample of eligible older adults.

### Research team and reflexivity

RA, a female physician and trained qualitative researcher, conducted the interviews. She had no prior relationship with the participants. Participant safety concerns were addressed with KL, the lead clinician. RA used reflexive notes to document her assumptions about how her age, gender, cultural identity, and professional background might affect participant interactions; her subjective responses to the interview context and interactions; post-interview reflections; and initial analytical memos. Research steps and decisions from project inception to the reporting of findings were recorded in an audit trail.

## RESULTS

### Participant characteristics

Interviews were conducted with 52 older adults: 16 White British, 20 South Asian and 16 African or Caribbean, with a mean age of 73 years and a majority of females (n=37; 71%). Ten interviews were conducted in non-English languages: Wolof (n=1), Luganda (n=2), Urdu (n=4), Punjabi (n=2) and Sylheti (n=1).

Although all participants self-reported current or past personal experiences of anxiety, it was during the interviews that the interviewer had the opportunity to gain a clear understanding of what the participants’ experiences of anxiety encompassed. At this stage, it was evident that our sample included two subgroups of participants: those who, at the time of experiencing anxiety, also self-reported experiencing significant distress (subjective experience of emotional pain communicated as an inability to cope with anxiety) and/or functional impairment (limitations in important areas of functioning caused by anxiety) and those who did not. Participants in the first group are referred to as those ‘with distressing anxiety’, while participants in the latter group are referred to as those ‘with non-distressing anxiety’. The data in this study incorporated the perspectives of 27 older adults with distressing anxiety and 25 with non-distressing anxiety. We established a typological classification to capture the differences in perspectives between the two groups of older adults within and across the three cultural groups. Participants’ characteristics are presented in **Appendix-3**

### Thematic overview

Participants’ beliefs about anxiety were aligned with the illness-related dimensions proposed in Leventhal’s Common-Sense Model: cognitive representation (identity, perceived causes, timeline, consequences, personal control/curability, coherence) and emotional representation. Inductive data analysis revealed two additional themes: perceived aggravating and protective factors.

#### 1) Identity

##### Label

Across all cultural groups, older adults who had used mental health services tended to be more comfortable using Western medical labels such as *‘anxiety’, ‘panic attacks’* and *‘depression’*. Conversely, those who had not accessed these services tended to avoid using medical labels. White British participants relied on colloquial terms such as *‘worrier’, ‘mither’, ‘stress’ and ‘worry’*, while South Asian and African/Caribbean participants used terms like ‘*pressure’*, *‘bothered’* as well as *‘worry’*, *‘stress’* and their synonyms in Non-English languages, or phrases like *‘Interpreter: I feel I have no peace’, ‘keyed up’, ‘the things I felt in my heart’* and *‘thinking a lot’*. South Asian and African/Caribbean participants noted how they avoid using specific native language labels when discussing their mental health state with others. In these groups, culture and religion were strongly intertwined. Thus, labels that implied non-conformity with cultural norms and denoted a *‘shameful’* lack of faith were avoided. Labels were also avoided if they indicated that one’s mental state had exceeded the community’s threshold of tolerance, thus inviting the substitution of empathy with control and intrusion by community members. In some cases, participants believed that the existing vocabulary could not capture their mental state, as available labels were exclusively reserved for more serious mental health problems.

> *‘Interpreter:…[Sylheti word: oshanti] it can be stigmatising, she’s saying, because in our culture some people will look at it, ‘Oh, she’s saying shame, it’s shameful,’ like, ‘Oh how could,’ you know, look down upon that feeling, people will think, you know, you haven’t got faith, why you’re feeling like this, you know, if you pray to God about it and you shouldn’t feel like that,….’ (South Asian female; distressing anxiety)*

> *‘Interpreter: It [Luganda word: okweraliikirira] doesn’t, it doesn’t bring a stigma per se, but what it does is that it gets everyone feeling sorry for you all the time, and they try to comfort you. But as much as they try to comfort you, they also get to a control point where they tell you, you need to change positions, you need to change feeling like that.’ (African/Caribbean female; distressing anxiety)*

Participants with non-distressing anxiety who had not accessed mental health services noted that the label *‘anxiety’* had recently *‘entered our language’* and was being *‘applied far more liberally’* in society. They suggested that this trend arose from unrealistic expectations of a stress-free life and that, at times, it may be exploited for secondary gains. Thus, *‘anxiety’* was considered a personally, socially and culturally unacceptable label.

> *‘I fear that it [anxiety] is this kind of thing that allows people to run away from living and getting on with life… I think it’s a title which is being applied for all kinds of things, which in my book you would get up, that’s what life’s about, that’s what living’s about.’ (White British male; non-distressing anxiety)*

> *‘…here if they have anxiety they believe that they can make money from the rich ((laughs)). I’m really sorry to say. Some people will say, “Oh because my boss talked to me like this, oh, now I’m stressed, oh now I have worriness, now I have anxiety.” (African/Caribbean male; non-distressing anxiety)*

##### Awareness of the Western label ‘anxiety’

Care was taken to use the same labels used by the participants during their interviews. However, all participants were asked about their understanding of the Western medical labels *‘anxiety’* and *‘depression’*. Most participants across all cultural groups recognised *‘depression’*, describing it as a *‘heavy’*, *‘very clearly definable illness’* that puts you *‘in a dark hole’*.

Non-English speakers recognised *‘depression’* and believed that people *‘Interpreter: get scared of the word depression’*, as it refers to a serious illness with negative consequences, *‘Interpreter:…some people commit suicide.’* Most of them could not find synonyms for *‘depression’* in their native language and used the English label instead.

Conversely, participants’ awareness of the Western label *‘anxiety’* differed across and within all cultural groups. Most White British participants identified *‘anxiety’* and *‘depression’* as distinct yet closely linked mental health conditions, with anxiety often tied to worry. However, this finding was not echoed in the South Asian and African/Caribbean groups where awareness of the label *‘anxiety’* varied widely. Those with healthcare backgrounds recognised *‘anxiety’*, believed that it was linked to worry, and identified it as distinct yet closely ‘*linked’* to depression. Although they found synonyms for anxiety in their languages, they believed that these synonyms did not convey the same severity as the medical terms *‘anxiety’* and *‘depression’*.

> *‘…[Hindi language: ‘ghabrana’, ‘ghabarāhaṭa’], in Hindi they will use this here. Anxious means anything that makes you very worried. You get [Hindi language: ‘ghabrana’]…They [people in home country] don’t take it as serious as we take it here [in the UK]. You know, if you say “anxiety”, it is in a medical term, yeah. If we say, “depression”, it is a medical term, isn’t it?’ (South Asian female; non-distressing anxiety)*

Nonetheless, most South Asian and African/Caribbean participants either believed *‘anxiety’* referred to a negative mental state distinct from depression, yet remained uncertain about its specific nature, or believed *‘anxiety’* and *‘depression’* referred to the same condition. This latter belief was especially true among participants whose native languages employed a single term for both.

> *‘I think it [anxiety and depression] comes under one umbrella, [Gujarati language: mansik], “mental health”, everything goes under that umbrella.’ (South Asian female; non-distressing anxiety)*

Additionally, a number of South Asian participants believed *‘anxiety’* referred to cognitive impairment or happiness.

> *‘Interpreter: She goes, anxiety…when you get older and you keep repeating yourself or repeating words.’ (South Asian female; non-distressing anxiety)*

> *‘Interpreter: She thinks “anxiety” is happiness and depression is upset.’ (South Asian female; distressing anxiety)*

##### Perceived symptoms

Across all cultural groups, participants with non-distressing anxiety largely considered symptoms of anxiety to be cognitive, whereas those with distressing anxiety experienced diverse cognitive, affective and somatic symptoms. Participants often found it challenging to discern if their symptoms, particularly somatic symptoms, were due to anxiety or comorbid mental and physical health problems. Notably, some South Asian participants expressed anxiety in terms of loss of interest and low mood.

#### 2) Perceived causes

Across all cultural groups, participants believed multiple causes contributed to anxiety. These causes differed across and within the cultural groups, with notable distinctions between participants with distressing and non-distressing anxiety. Participants with distressing anxiety tended to rely on their personal experiences when recounting causes of anxiety, which mainly were unmodifiable stressors that changed across their lifespan. An enduring sense of cumulative losses permeated these accounts - loss of health, loss of independence, loss of companionship, loss of sense of value, security or belonging. Conversely, participants with non-distressing anxiety tended to cite more general causes. We classified causes of anxiety into three categories: intrapersonal, interpersonal and external.

Intrapersonal factors were the most frequently cited across all cultural groups. However, there were marked differences between participants with distressing and non-distressing anxiety. Those with distressing anxiety primarily attributed anxiety to declining physical health and the subsequent loss of independence. They often reflected on their lived experiences of progressive health issues that rendered them constantly ruminating about who they once were and worrying about who they would become.

> *‘So my anxiety in fact wasn’t bad initially but as the symptoms got worse I suppose I was anxious…now I’m more anxious all of the time because my Parkinson’s is so much worse…I mean, I hate, it makes me anxious when people see you as an invalid or somebody who’s disabled. But you can’t mistake it now. It’s obvious that I have Parkinson’s now.’ (White British female; distressing anxiety)*

These participants grappled to understand why they acquired these health issues and found it challenging to accept their detrimental impacts, *‘I just thought, why did it have to affect me, you know, why me, you know?’* While they considered anxiety to be a justifiable emotional response, inextricable from their experience of physical illness, they did not perceive it as a normal consequence of ageing. They were left in limbo - a state of uncertainty. In contrast, those with non-distressing anxiety, typically with well-controlled physical health problems, tended to view anxiety as an expected *‘ageing thing’*.

Participants with non-distressing anxiety identified negative personality attributes that incline individuals to *‘think very negatively all the time’* as the primary cause of anxiety. People with these traits were considered *‘weak’*, *‘moaner(s)’* who could not cope with stressors and were held responsible for their negative *‘mental attitude*’. Only South Asian and White British participants who had experienced distressing anxiety since childhood and could not identify other possible causes for anxiety believed anxiety was part of their *‘chemical build-up’*, *‘it’s part of who I am’*.

Lack of faith was the only culture-specific intrapersonal cause mentioned exclusively by South Asian and African/Caribbean participants with non-distressing anxiety. These participants tended to have salient religious identities and believed anxiety was an indication of failure to cope with stressors using religious means.

> *‘Interpreter: Because the devil makes us to get worried…What I would say about people who worry is that they don’t have God or they don’t believe, because even Jesus told us not to worry. Instead of worrying, pray in all circumstances, pray, pray.’ (African/Caribbean female; non-distressing anxiety)*

Interpersonal causes included both shared cross-cultural and distinct culture-specific social losses. Participants from all cultural groups identified family issues, loneliness and loss of valued roles as causes of anxiety. However, cultural norms and expectations introduced additional layers of complexity, particularly among South Asian and African/Caribbean participants with salient cultural or religious identities.

While worrying about one’s children was shared across all groups, South Asian participants were explicitly concerned about their children living outside their community and the *‘lack of control’* over a generation that may not respect its *‘elders’*. Additionally, grown-up children were expected to abide by cultural norms and care for their parents. Hence, disruptions in family unity by an *‘in-law’* wanting to ‘*get separated from the family’* was regarded as an important cause of anxiety.

> *‘Interpreter: Because obviously when [participant’s name] was married back home, she lived with her mother-in-law, looked after the family and she never felt like she was, you know, you’re being used. But now that she’s got her kids married, they’ve married into modern relationships where it’s about their, yeah, so they just don’t want to look after her or even want to just come and visit so they’re helping her mentality, you know, mental health.’ (South Asian female; distressing anxiety)*

Marital problems were cited as a cause of anxiety across all cultural groups. African/Caribbean participants mentioned separation but tended to avoid unpacking their experiences. Conversely, White British and South Asian women described how being stripped of control by their *‘controlling and domineering’* husbands contributed to their mental health problems. However, these experiences were more conspicuous among some of the South Asian women who experienced verbal and physical abuse and strict control by their husbands’ families, with cultural norms intensifying their sense of entrapment and anxiety.

> *‘…I would have to put up with all this, and that’s when my anxiety and depression kicked in. So then, my parents were good, but they would say, “Well, you’ve got a roof over your head, and he gives you money for food, what you’re going to do, you’ve got four children?’’ (South Asian female; non-distressing anxiety)*

Caring for a family member with declining physical or mental functioning was a significant precursor of anxiety across all cultural groups. Participants felt that caring for their loved ones ‘properly’ was a moral obligation and were thus constantly alert. This obligation commanded that they manage and even give up competing responsibilities, *‘I still deprived everything and looked after her [participant’s mother].’* South Asian and African/Caribbean participants, in particular, felt overwhelmed by the added pressure of meeting other family members’ expectations regarding caregiving.

> *‘…everybody’s ringing me up, can you do this, can you do that, and sometimes, they forget, I think they forget about me sometimes as well, you know how I’m coping.’ (African/Caribbean female; distressing anxiety)*

While adopting the caregiver role constituted a significant shift in one’s valued roles, participants also struggled with losing other valued roles. They lost parents, partners, children and siblings and believed that *‘Bereavement is another form of stress.’* That sense of loss was amplified among South Asian and African/Caribbean participants with salient cultural identities when they were deprived of sharing their grief with or finding adequate support from members of their communities.

> *‘…the hardest part of it was in our culture, any family relatives pass, our community they would visit you, give you their support, and just sit with you for a little while and talk, if you want to talk through things. But at the time my daughter passed, the lockdown came and no one could visit me. There was the phone so they could contact me via the phone, but that visit, that face to face, that culture thing wasn’t there at all…’ (African/Caribbean female; distressing anxiety)*

Retirement constituted another form of loss described by White British and African/Caribbean participants with distressing anxiety as a loss of one’s longtime personal goals, sense of value and identity, *‘I feel like sometimes I have left my identity, lost my identity.’*

These cumulative social losses led to loneliness - a shared cross-cultural cause of anxiety. However, it was predominantly South Asian and African/Caribbean participants who emphasised the role of loneliness and lack of quality family support in anxiety. Loneliness meant more time for *‘thinking so much’* and not finding a caring, trustworthy person to talk to, *‘If people had someone to talk to the psychiatric wards would be empty.’* They accused individualistic Western communities of promoting loneliness.

Cultural group-specific interpersonal causes of anxiety included adverse childhood experiences and upward social comparison. Several White British and South Asian participants attributed anxiety to traumatic childhood experiences, including emotional, physical and sexual abuse and living during an era where *‘you are being persecuted’,* such as World War Two. Participants with distressing anxiety described how these experiences impacted their sense of security, agency and self-worth, resulting in complex mental health problems and suicide attempts.

South Asian and African/Caribbean participants with non-distressing anxiety believed that comparing oneself with others, perceived to be superior in terms of materialistic possessions, resulted in anxiety. Upward social comparison was interpreted as a sign of lack of faith since it indicated a lack of gratitude for God’s blessings.

External causes of anxiety were exclusively cited by South Asian and African/Caribbean participants. These included workplace discrimination, asylum-seeking and forced displacement, and financial problems. Discrimination created a sense of alienation, *‘otherness’* and a loss of sense of value, belonging and security, especially when there were no means of protection.

> *‘So the look, is the systematic racism that it is very hard to pinpoint it. If you do anything, if you walk and you don’t say thank you, you get reported. “Oh, she has done this. Oh, she did this.” And you can’t defend yourself because you are alone. The majority carries the vote. So it is like any day you are going to work, you don’t know who you are going to work with, you don’t know what mess will be coming up. You live in tension.’ (African/Caribbean female; distressing anxiety)*

Similarly, participants who were forced to leave their home countries and seek asylum in other countries explained how they had to leave behind meaningful social networks, their own culture, traditions and language - their sense of belonging, *‘Interpreter: So it’s like picking up a bird and totally going and putting it into a different, unknown location.’* In some cases, participants’ anxiety was exacerbated by the uncertain, complex and lingering nature of the asylum-seeking process.

Participants recognised *‘the cost of living’* as a precursor for anxiety. However, those who experienced distressing anxiety tended to recount the devastating impact financial difficulties had on their lives and mental health.

> *‘Interpreter:…he [participant’s son] lost his child and, if I had had money maybe my grandchild could have survived.’ (African/Caribbean female; distressing anxiety)*

Figure 1 illustrates the cross-cultural comparison of the perceived causes of anxiety.

**Figure 1:**
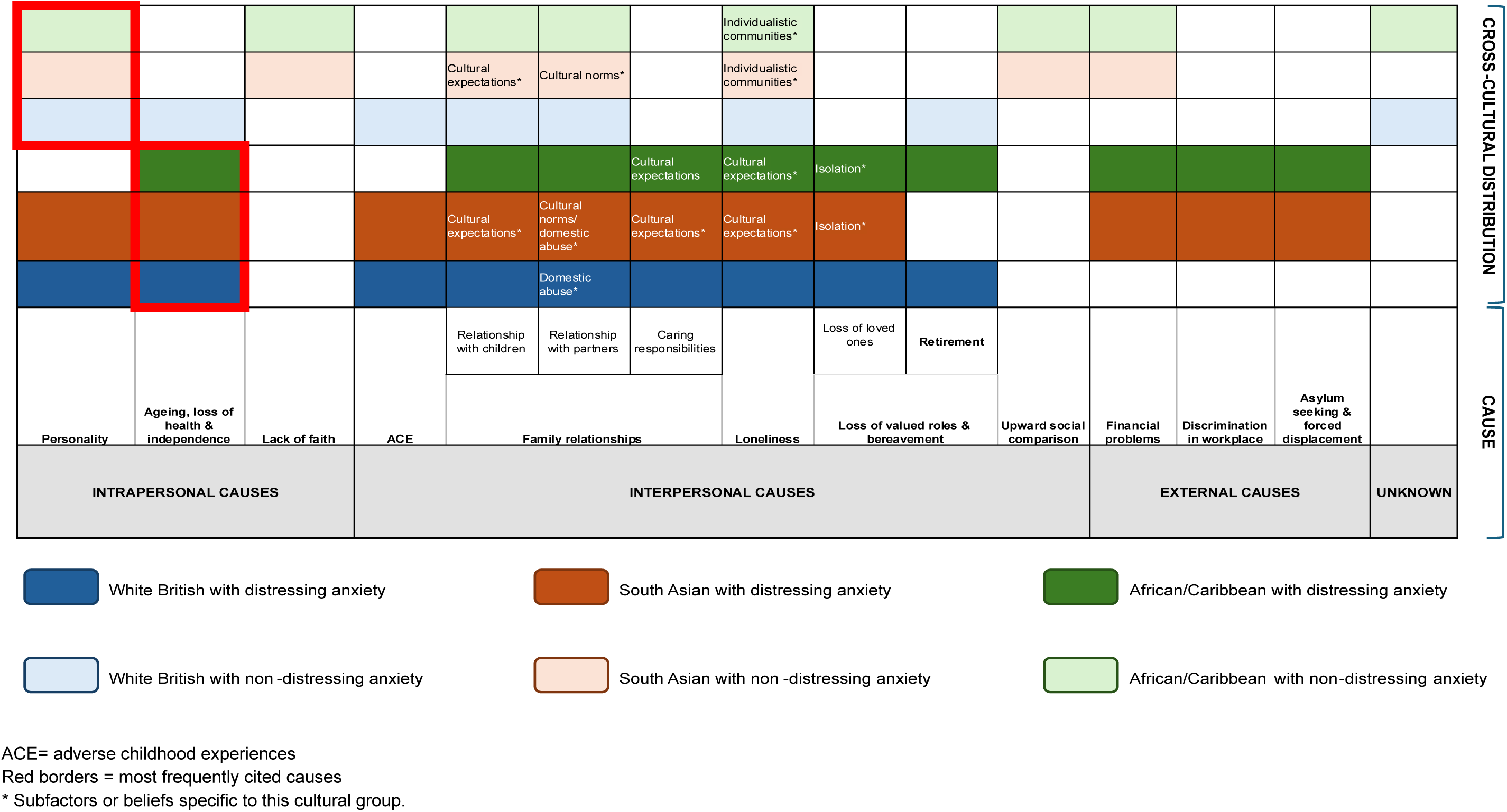
Cross-cultural comparison of perceived causes of anxiety

#### 3) Perceived course

Participants identified the onset of anxiety as the time in their lives or the event after which they experienced symptoms of anxiety: during childhood, early adulthood or later in life. In some cases, this transition was noticed by the participant, while in other cases, it was noticed by others, such as family members or general practitioners.

When asked about the course of anxiety, participants with distressing anxiety offered a range of responses based on their personal experiences and the current stage in their illness journey. Most participants described anxiety as a chronic condition with either a progressive, static or fluctuating course. Participants who experienced what they believed were panic attacks described anxiety as an acute, recurrent state. Participants who believed they had reached a stage in their illness journey where they had successfully coped with anxiety viewed it as a chronic condition with a regressive course.

Conversely, participants with non-distressing anxiety tended to describe anxiety as an acute, short-term emotional response to different stressors, *‘…it [anxiety] comes and goes, it [anxiety] is not continuous.’*

#### 4) Perceived controllability/curability

Participants’ views regarding the curability and controllability of anxiety varied widely according to their focus of control. No specific cross-cultural patterns were identified. Participants who focused solely on unmodifiable causes of anxiety and their inability to eliminate them believed anxiety was an incurable, uncontrollable condition.

> *‘Well, it’s not actually saying, I haven’t got enough bread, I deal with it by buying some bread. I can’t deal with it [anxiety], I can’t make it [anxiety] go away,…’ (White British female; distressing anxiety)*

Participants who believed causes of anxiety were unmodifiable, but shifted their focus on coping strategies they could use to control anxiety symptoms believed it was an incurable, but controllable condition.

> ‘*…you can control it [anxiety], and I control it, I like to think, by the two ways of exercise and the distraction techniques, but actually curing it, I just don’t know if there is such a thing as a cure.’ (White British male; distressing anxiety)*

In contrast, most participants with non-distressing anxiety believed anxiety was a self-manageable condition that could be ‘*easily overcome’* by enforcing simple strategies, such as ‘*avoiding the situations that make you anxious*.’

#### 5) Perceived consequences

Most participants who differentiated anxiety from depression believed that failure to cope with anxiety could lead to more serious mental health problems, namely depression. Participants with distressing anxiety experienced significant mental and physical health ramifications, including alcohol misuse and the exacerbation of existing physical health problems. They described how anxiety adversely affected their social and occupational performance, thus worsening their circumstances and aggravating their mental health problems further.

> *‘…when I was in my teens, I started abusing alcohol, because I felt like it relaxed me, and over the years it developed into full blown alcoholism…I just realised what it cost me over the years.’ (White British male; distressing anxiety)*

#### 6) Aggravating factors

Aggravating factors were factors believed to trigger or worsen anxiety symptoms, but were not considered to be causes of anxiety. Although these included usual activities like driving or going to the supermarket, participants perceived them as threatening, assuming they could lead to unfavourable outcomes. Consequently, they were avoided to mitigate anxiety.

> *‘I love to go to the gym, but now, I’m scared nowadays because of my health. Because my knees are not working properly, so I’m scared if I go, maybe I fell down and in the old age, when you fell down, you broke your hip or leg or arm, and maybe it won’t recover because I’m diabetic as well so I limited my activities and I don’t want to go out.’ (South Asian female; distressing anxiety)*

#### 7) Protective factors

Protective factors were factors believed to relieve pre-existing mental health problems or prevent their occurrence in the first place. South Asian and African/Caribbean participants with salient cultural and religious identities regarded membership in communities aligned with their identities as protective factors against anxiety. Culture and faith were intertwined. Belonging to a group with whom one identifies on many levels gave participants *‘a sense of belonging’* and support. Additionally, those who immigrated in adulthood believed that prior exposure to socioeconomic hardship built their resilience and made them more equipped to manage anxiety.

> *‘So my faith is that, in the Church we behave like brothers and sisters. My pain is their pain…When you meet together you hear each other’s worrying’s, you relieve it. But when you buckle it alone, it’ll send you to these mental things, the Alzheimer’s, the dementia, the confusion, call it any psychiatric disorder. So faith is a very important thing to have. But those who don’t have faith I don’t know how they live…’ (African/Caribbean female; distressing anxiety)*

#### 8) Emotional representations and perceived incoherence of certain aspects of illness beliefs about anxiety

Participants with distressing anxiety experienced psychological distress, shame and worry. Across all cultural groups, psychological distress was usually communicated as emotional pain resulting from perceived functional impairment and the inability to cope with anxiety - *‘helplessness’*.

> *‘I feel I’m a weak person, I’m hopeless. I feel useless at times.’ (South Asian female; distressing anxiety)*

The findings also suggest that confusion about certain aspects of anxiety may have contributed to a sense of shame. White British participants who could not identify external triggers for anxiety and experienced generalised worries about what they believed were *‘trivial’, ‘stupid little things’*, felt *‘silly’* and refused to discuss it with others. South Asian participants with salient religious identities were conflicted by their experience of distressing anxiety. One participant explained how she had *‘strong willpower’* and believed in the power of her faith, but at the same time, she believed anxiety was for the *‘very weak’* who had *‘no willpower to tackle these things’.* Participants also expressed concerns about being worried about their anxiety and the repercussions of this negative feedback loop*, ‘I was worried about it [anxiety] and it causes more worry about it.’*

#### 9) Comparison between anxiety and depression

Across all cultural groups, those who distinguished between anxiety and depression believed depression was ‘*a much deeper’,* ‘*more permanent’*, *‘clinical thing’* that was *‘harder to deal with than anxiety’* and necessitated treatment. Participants attributed depression to both biological - *‘a chemical that’s missing’* due to chronic medical health problems, *‘brain weakness’* or a *‘genetic’* problem that runs in families – and social causes. They also believed it could be a consequence of continuous negative thinking and anxiety.

## DISCUSSION

### Summary of the findings

This multicultural qualitative study aimed to explore and compare beliefs about anxiety among self-reporting White British, South Asian and African or Caribbean older adults living in the UK, informed by Leventhal’s Common-Sense Model. The study incorporated the perspectives of 27 older adults with distressing anxiety and 25 with non-distressing anxiety. Overall, participants’ beliefs about anxiety mapped onto the a priori illness-related dimensions proposed by Leventhal’s Common-Sense Model. The findings reveal how beliefs about anxiety differed across and within all cultural groups, with notable distinctions between participants with distressing and non-distressing anxiety. Through inductive analysis, we identified two new dimensions: aggravating and protective factors. People’s salient identities, rather than their cultural background, shaped their understanding of anxiety.

### Special aspects of older adults’ beliefs about anxiety

Recognising an illness or health threat is the first step in managing it. Previous reviews indicate that older adults often normalise depression (21, 22) and anxiety (7) as typical responses to ageing-related comorbidities, functional impairment and social stressors. In our study, this was only true for participants with non-distressing anxiety. In contrast, participants with distressing anxiety viewed it as a justified emotional response to uncontrollable stressors; however, one that exceeded what they considered as a ‘normal’ mental state, caused significant distress and impairment, left them feeling *‘helpless’* and unable to cope. These participants were left in limbo; they neither viewed anxiety as a ‘normal’ part of ageing nor considered it as an ‘illness’ trajectory.

This ambiguity in distinguishing ‘normative’ adaptive anxiety from ‘non-normative’ pathological anxiety (i.e., anxiety disorders) reflects a long-standing dilemma (23), which appears to have perplexed older adults in our study. Leventhal’s Model posits that perceived deviations from the ‘normative’ self initiate self-regulatory processes (24). However, while the subjective boundary between ‘normative’ and ‘non-normative’ states may appear relatively clear when experiencing overtly abnormal symptoms, like seizures, it becomes far more ambiguous when symptoms overlap with what is considered normal human functioning. Both anxiety and fear fall within the spectrum of normal emotional responses (25). Therefore, symptoms which constitute the diagnostic criteria for ‘non-normative’ anxiety are widely and variably distributed throughout the population; existing along a continuum rather than as an all-or-none phenomenon (26). This raises an important question: if people conceptualise ‘illness’ as a dichotomous entity, how do they determine the boundary between ‘illness’ and ‘normality’ within the inherently dimensional nature of mental disorders? This mismatch may partly explain why older adults struggled to classify their distressing anxiety as an ‘illness’, despite recognising it as abnormal. Moreover, perceiving distressing anxiety as rooted in social suffering may further impede its classification as an ‘illness’ - a trajectory typically framed within biomedical explanations. The heterogeneity of anxiety disorders (27), and the high prevalence of comorbidities in this population pose further complexity, making it challenging to distinguish between disorders with overlapping symptoms (28). Together, these challenges blur the boundaries between ‘normality’, ‘abnormality’ and comorbidity in mental health. Therefore, it could be argued that Leventhal’s Model, a model originally developed for physical health, may be ill-equipped to capture the nuanced complexity of symptom appraisal in mental health contexts.

Researchers have also argued that the encroachment of mental illness upon the self poses a conceptual challenge to Leventhal’s Model, which requires a distinct, unaffected ‘self’ to appraise the ‘illness’ (29). Nonetheless, it is important to acknowledge that, much like mental illness, the boundary between ‘illness’ and ‘self’ is similarly blurred in chronic physical conditions, where individuals reconstruct their identity in response to illness along a continuum, from accepting the illness as part of the self to experiencing it as engulfing the self (30).

Furthermore, our findings indicate that although older adults’ understanding of anxiety was often unclear, their beliefs largely aligned with the dimensions posited in Leventhal’s Model, supporting the model’s applicability to anxiety in older adults and underscoring its broader relevance to mental health. Moreover, the model’s applicability across both physical and mental health domains highlights the need to move beyond the contrived divide between physical and mental health problems, particularly in populations at high risk of comorbidity. Chronic physical health problems are common in the older adult population, and understanding an individual’s beliefs about mental health concerns such as anxiety and depression cannot occur in isolation of their beliefs about other comorbidities, their impact on mental health and the intricate ways in which coexisting disorders interact. This complexity is highlighted in our findings which show that older adults with distressing anxiety attributed it primarily to progressive health issues, declining physical health and loss of independence, rendering anxiety and physical illness inextricably linked.

Older adults’ awareness of the labels ‘anxiety’ and ‘depression’ provided further insights. Most participants, including non-English speakers, recognised the label ‘depression’, but not ‘anxiety’. Depression was usually referred to using the Western label, while anxiety was referred to using synonyms of stress, *‘pressure’* or *‘thinking a lot’*. In some languages, both conditions were merged under one term; in others, labels were reserved for more severe problems. This finding is a reminder that these diagnostic labels “constitute a folk taxonomy” (31), and that, distinctions between ‘anxiety’ and ‘depression’ while meaningful in English may not be so across other languages. In these languages, anxiety and depression may be combined into a broader category of emotional distress given the overlap of symptomatology, high rates of comorbidity and the common predominance of social explanatory models.

Labels also reflected what participants perceived as personally, socially and culturally acceptable mental states. Those who accessed mental health services were more accepting of Western medical labels. Others, particularly those with salient religious or cultural identities, mainly in the South Asian and African/Caribbean groups, refuted medical labels that implied weak faith or unacceptable deviation from cultural norms. However, these views were not universal within the South Asian and African/Caribbean groups, but rather held by those who strongly identified with their cultural or religious groups.

The impact of people’s identities on beliefs about anxiety was also emphasised when discussing protective factors. South Asian and African/Caribbean participants with salient religious and cultural identities viewed these social identities as buffers against mental health problems. Nevertheless, the impact of social identities on older adults’ mental health may not be as straightforward. Examining the perceived causes of anxiety reveals that adhering to cultural norms may have aggravated mental health concerns among some of the South Asian and African/Caribbean participants. These seemingly contradictory effects of social identification reflect the complexity of social identities which vary along a number of dimensions: their relative importance, appraisal and the degree to which the individual is attached to and immersed within the group (32). Moreover, people possess multiple personal (unique, idiosyncratic characteristics), role (e.g., caregiver), and social identities (e.g., culture/religion) (33), which intersect, interact and influence each other. The impact of these multiple identities on one’s well-being depends on the number of endorsed identities, their salience, distinctiveness and potential conflicts between them (34).

Our findings caution against grouping people into broad, arguably homogeneous entities of discrete cultural groups. Doing so overlooks the variable influence of culture within groups, the complexity and diversity of individual experiences (35), and the different ways individuals construct their identities and want to be perceived by others. This can lead to stereotyping, which is not only inaccurate, but can be potentially harmful (36).

## STRENGTHS AND LIMITATIONS

This study represents the first multicultural qualitative study to explore older adults’ beliefs about anxiety. Key limitations include recruiting a self-reporting sample and using different terms to describe anxiety. Our public contributors were concerned that using Western medical labels could suggest that only individuals with a clinical diagnosis of anxiety disorders or those who had used mental health services were eligible to participate, thus excluding potentially eligible older adults with undiagnosed anxiety. That is a highly relevant group for whom studies on anxiety perceptions and coping strategies are lacking. Public contributors who self-identified as South Asian or Caribbean were concerned that Western medical labels may not be easily recognisable in their communities and, if recognised, may have stigmatising connotations. These labels were also difficult to translate into other languages, which is problematic when attempting to reach non-English speakers. Therefore, we decided to include lay terms such as “worrier” and “stress” in the study adverts. However, using these terms may have allowed the inclusion of participants with a wide range of experiences, thus introducing variability in the data. Although several measures were taken to ensure participants included in the study experienced anxiety (current or past), we believe that the sample included participants who could probably fulfil the diagnostic criteria for anxiety disorders (i.e., those with distressing anxiety) and those who could not (i.e., those with non-distressing anxiety). We established a typology to highlight the differences between both groups. Additionally, working with different interpreters to facilitate interviews with non-English-speaking participants could have impacted interpretation consistency.

## CONCLUSIONS

The current study has built on previous research confirming the applicability of Leventhal’s Model in mental health contexts. Our findings suggest that older adults tend to have a fragmented understanding of anxiety. Participants with distressing anxiety did not normalise anxiety, nor did they recognise it as an illness trajectory. Notably, specific aspects of older adults’ beliefs about anxiety were shaped by their salient identities rather than their cultural backgrounds. Grouping people into broad, arguably homogeneous entities of discrete cultural groups disregards the diversity and heterogeneity of individuals within the same cultural group. Cross-cultural research should embrace this diversity and adopt more nuanced approaches to provide meaningful, respectful and person-centred insights into people’s understandings and experiences of illness.

## AUTHOR CONTRIBUTIONS

Rasha Alkholy (RA) contributed to funding acquisition, project management, study conceptualisation, methodology, investigation, data curation, formal analysis, visualisation, writing- original draft and preparation.

Karina Lovell (KL), Rebecca Pedley (RP) and Penny Bee (PB) provided supervision and guidance throughout the whole research process, contributed to investigation (cross-checking data analysis and interpretation) and review and editing of the publication drafts.

## ACKNOWLEDGEMENTS

We would like to express our sincere gratitude to the participants who generously gave their time and shared their experiences, rendering this research possible. We are extremely grateful to our public contributors for their collaboration on this project from inception. We extend our thanks to the community and charity organisations that hosted our arts-based workshops and presentations, and those that helped us reach a wider audience of potential participants: Older Black Africans Day Opportunities (OBADO), Khush Amdid, Yellowbird Age Friendly, Hulme Carer’s Forum, African Caribbean Care Group, Parkinson’s UK, Stroke Association Carers Group, Growing Old Disgracefully, GM Older People’s Network and Dementia United.

## FUNDING DETAILS

This work was supported by the President’s Doctoral Scholar Award (a PhD studentship awarded by The University of Manchester, United Kingdom). The funder had no role in the study design, collection, analysis or interpretation of the data, writing the manuscript, or the decision to submit the paper for publication.

## DATA AVAILABILITY

The anonymised dataset is available for restricted data-sharing to authenticated researchers who provide verifiable institutional affiliation and have ethical approval in place. A metadata record of the anonymised dataset is available on Figshare (https://doi.org/10.48420/28468466) for further information and guidance.

## DISCLOSURE STATEMENT

The authors confirm that there are no relevant financial or non-financial competing interests to report.

### Appendix 1: Public contributor commentary

Authors (authorship details in accordance with individual preferences):

A, Lewis; A, P; F, Greenmantle; M, Godden; N, Rehman

This cross-cultural study is the first of its kind and is really important. It highlighted how people from different communities used and understood labels of anxiety. In addition, it raised awareness of anxiety in people’s minds. Some people never had the opportunity to talk about their feelings and this research provided them the opportunity to acknowledge and validate those feelings and offered them a safe space to talk openly about their feelings without judgement. Everyone in our group has had a personal experience of anxiety. Therefore, we find this of relevance to ourselves.

Discussions about study findings that we found surprising as a group were insightful. Although we expected to find differences between the cultural groups in our study, it was also interesting to find differences among our own group of public contributors. Some White British public contributors among us were surprised about the lack of reliance on God as a reason for anxiety. They understood that some cultures do not have a word for anxiety, but to put it down as a failure in yourself to believe in God was surprising. In contrast, the South Asian public contributors among us were not surprised by this finding because they believed that faith was an integral part of the South Asian identity. In their view, South Asian parents and the community at large indoctrinate children into cultural beliefs, which are often influenced by religious scripture, as they grow up. These beliefs and teachings stop people from sharing feelings openly with others for fear of judgement or the desire for acceptance from other community members. Similarities across cultural groups were interesting as well as some of us were not expecting them. An important finding that echoed our own experiences was that ageing and the experiences one has when getting older can be very stressful, for example, bereavement. From our personal experience, retirement may not have gone as expected, especially if early retirement was a forced decision due to ill health. This resulted in a loss of identity and caused anxiety and depression.

Giving feedback to the communities involved in this study is important because it will allow them to use the study findings to support their own communities. It will ensure that their participation is not tokenistic. We also need funding to raise awareness about anxiety in older adults, tell people what anxiety is, its impact on their lives and inform them of available support. We believe that different service providers, including the British Association for Counselling and Psychotherapy, the therapeutic profession, GPs, medical professionals and organisations dealing with diversity should be made aware of this research. Community organisations working with older adults must be involved in future research because they contribute far more positive therapeutic counselling than is recognised at the moment.

Because of COVID-19, the time scale for the study was extended. The original plan was to meet face-to-face, but Zoom meetings were an adaptation to the plan. When we first started doing Zoom meetings, we were not used to them; managing large numbers of people at a Zoom meeting can sometimes be disruptive, we cannot read physical cues and body language. Because of funding limitations on the study, we were restricted to three cultural groups. Possible future research could build on this and definitely include more cultural groups, and possibly larger numbers of participants. More research is required in this field.

This is a ground-breaking study dealing with anxiety and culture. It highlights the way different cultures deal with and think about anxiety. We are hoping it will act as a starting point from which service providers and community organisations can develop and deliver culturally appropriate support. We believe that as a group we have gained new skills and knowledge about research in general, and anxiety and depression in particular. Being involved in this research made us feel valued and gave us the opportunity to meet new people.

## Appendix 2: Semi-structured interview topic guide

|  |  |
| --- | --- |
| **Appointment details** |  |
| **Participant Identification Number (PIN)** |  |
| **Date** |  |
| **Time** |  |
| **Interview type** | ☐ Phone<br>☐ Zoom |
| **Language used during the interview** |  |
| **If the interview is to be conducted in a non-English language, please complete the following:** |  |
| • **Language used** |  |
| • **Name of the interpreting services supplier used** |  |
| • **Did the linguist give verbal consent to audio-record this interview?** | YES / NO |

| **1. Experience and understanding of anxiety** |  |
| --- | --- |
| **Objective** | To explore illness beliefs of anxiety from the perspective of community- dwelling older adults with anxiety. |
| **Probes/ Prompts** | • **Explore participant's own experience and understanding of anxiety:**<br>○ Word(s) used to describe anxiety/words used by others<br>○ Emotions or feelings<br>○ Thoughts<br>○ Symptoms (bodily symptoms)<br>○ Duration, timeline<br>○ Severity<br>○ Causes<br>○ Effect on life (consequences) |
| **2. Coping strategies** |  |
| **Objective** | To explore older adults' choice of coping strategies in response to anxiety. |
| **Probes/**<br>**Prompts** | • **Choice of coping strategies:**<br>○ Reasons for choosing that strategy<br>○ Perceived effectiveness |
| **3. Help-seeking behaviour** |  |
| **Objective** | To explore older adults' help seeking behaviour in response to anxiety. |
| **Probes/**<br>**Prompts** | • **Previous experience of seeking advice from formal mental health services (e.g. GP, psychologist, etc.):**<br>○ From whom<br>○ Strategy used to manage anxiety<br>○ Perceived effectiveness<br>• **If the participant has not previously sought advice, explore their views about asking for advice from formal mental health services:**<br>○ From whom<br>○ Views about when (and why) it is appropriate to ask for advice<br>• **Talking about 'anxiety' to your GP:**<br>○ Difference in the way anxiety is described to GP compared to others, and why |
| **4. Barriers and facilitators to mental health services utilisation** |  |
| **Objective** | To explore barriers and facilitators to mental health services utilisation from the perspective of community-dwelling older adults with anxiety. |
| **Probes/**<br>**Prompts** | • **Factors that help encourage participants to use mental health services**<br>○ Views about facilitators<br>• **Factors that act as a deterrent to participants using mental health services**<br>○ Views about barriers<br>• **Awareness of different strategies available for managing anxiety**<br>○ Different strategies<br>○ Personal experience<br>○ Views about effectiveness<br>○ Views about advantages and disadvantages<br>○ Preference of a strategy over the other and why |
| **5. Outcome(s) of interest** |  |
| **Objective** | To explore outcome(s) of interest, from the perspective of community- dwelling older adults with anxiety, that are used to appraise coping strategies for anxiety. |
| **Probes/**<br>**Prompts** | • **Outcome(s) of interest used to appraise the success of management** |
| **Closing** |  |
|  | Any other issues that the participant would like to raise |

## Appendix 3: Participant characteristics

| Participant characteristic | White British | South Asian | African/Caribbean | Total |
| --- | --- | --- | --- | --- |
| Age (years), mean (SD) | 73.56 (6.57) | 70.85 (6.62) | 75.37 (8.37) | 73.08 (7.30) |
| Age range (years) | 66 - 91 | 65 - 86 | 66 - 88 | 65 - 91 |
| Female, n (%) | 10 (62.50) | 14 (70.00) | 13 (81.25) | 37 (71.15) |
| **Language used** |  |  |  |  |
| • English | 16 | 13 | 13 | 42 |
| • Non-English | 0 | 7 | 3 | 10 |
| **Living status** |  |  |  |  |
| • Living alone | 4 | 4 | 9 | 17 |
| • Living with others | 12 | 16 | 7 | 35 |
| **Needs help in performing daily activities** |  |  |  |  |
| • Always/most of the time | 3 | 6 | 1 | 10 |
| • Sometimes | 2 | 2 | 1 | 5 |
| • Rarely/never | 11 | 12 | 14 | 37 |
| **Medical problems** |  |  |  |  |
| • Yes | 16 | 18 | 13 | 47 |
| **Participants with distressing anxiety \*** | **9** | **10** | **8** | **27** |
| Currently experiencing symptoms suggestive of anxiety <sup>a</sup> |  |  |  |  |
| • Yes | 8 | 9 | 6 | 23 |
| Currently experiencing symptoms of depression <sup>b</sup> |  |  |  |  |
| • Suggestive of depression | 5 | 2 | 2 | 9 |
| • Almost always depression | 2 | 5 | 0 | 7 |
| Used health services for mental health problems |  |  |  |  |
| • Yes | 9 | 6 | 6 | 21 |
| Clinical diagnosis of mental health problems |  |  |  |  |
| • Anxiety | 1 | 1 | 0 | 2 |
| • Depression | 1 | 1 | 0 | 2 |
| • Anxiety and other comorbidities (depression, PTSD) | 3 | 2 | 1 | 6 |
| **Participants with non-distressing anxiety \*\*** | **7** | **10** | **8** | **25** |
| Currently experiencing symptoms suggestive of anxiety <sup>a</sup> |  |  |  |  |
| • Yes | 6 | 3 | 2 | 11 |
| Currently experiencing symptoms of depression <sup>b</sup> |  |  |  |  |
| • No depression | 7 | 10 | 8 | 25 |
| Used health services for mental health problems |  |  |  |  |
| • Yes | 2 | 3 | 2 | 7 |
| Clinical diagnosis of mental health problems |  |  |  |  |
| • Anxiety and other comorbidities (depression) | 0 | 0 | 1 | 1 |
| • Other mental health problems (e.g., bipolar disorder, OCD) | 0 | 2 | 0 | 2 |

\*Participants who self-reported current or past anxiety that was associated with distress &/or functional impairment.
\*\*Participants who self-reported current or past anxiety that was not associated with distress &/or functional impairment.
<sup>a</sup> Geriatric Anxiety Scale total score > 9: currently experiencing symptoms suggestive of anxiety; ≤ 9: currently not experiencing symptoms suggestive of anxiety (Gould et al., 2014).
<sup>b</sup> Geriatric Depression Scale-Short Form ≤5: no depression; 6-10: suggestive of depression; ≥ 11: almost always depression (Yesavage and Sheikh, 1986).
n=number; OCD=obsessive compulsive disorder; PTSD=post-traumatic stress disorder; SD=standard deviation.

## Notes

### Competing Interest Statement

The authors have declared no competing interest.

### Author Declarations

The Research Ethics Committee of The University of Manchester gave ethical approval for this work (Ref: 2021-10461-17766; Date: 05/02/2021).

## REFERENCES

1. Bryant C, Jackson H, Ames D. The prevalence of anxiety in older adults: Methodological issues and a review of the literature. J Affect Disord. 2008; 109(3): 233–250 10.1016/j.jad.2007.11.008.

2. Chen JTH, Belcher J, Zagic D, Wuthrich VM. Anxiety disorders in later life. In: Asmundson GJG, Pachana NA, (eds.) Comprehensive clinical psychology. 2nd ed. Oxford: Elsevier; 2022. p. 144–160. 10.1016/B978-0-12-818697-8.00020-0.

3. Wolitzky-Taylor KB, Castriotta N, Lenze EJ, Stanley MA, Craske MG. Anxiety disorders in older adults: A comprehensive review. Depress Anxiety. 2010; 27(2): 190–211 10.1002/da.20653.

4. Hohls JK, Konig HH, Raynik YI, Hajek A. A systematic review of the association of anxiety with health care utilization and costs in people aged 65 years and older. J Affect Disord. 2018; 232: 163–176 10.1016/j.jad.2018.02.011.

5. Byers AL, Arean PA, Yaffe K. Low use of mental health services among older Americans with mood and anxiety disorders. Psychiatr Serv. 2012; 63(1): 66–72 10.1176/appi.ps.201100121.

6. Mackenzie CS, Reynolds K, Cairney J, Streiner DL, Sareen J. Disorder-specific mental health service use for mood and anxiety disorders: Associations with age, sex, and psychiatric comorbidity. Depress Anxiety. 2012; 29(3): 234–242 10.1002/da.20911.

7. Alkholy R, Lovell K, Bee P, Pedley R. Barriers and enablers to help-seeking behaviour for mental health reasons among community-dwelling older adults with anxiety: Mixed-methods systematic review. Journal of Affective Disorders Reports. 2022; 10(100440): 10.48420/28468466.v2.

8. Kirmayer LJ, Bhugra D. Culture and mental illness: Social context and explanatory models. In: Salloum IM, Mezzich JE, (eds.) Psychiatric diagnosis: Challenges and prospects. John Wiley & Sons, Ltd; 2009. p. 29–40. 10.1002/9780470743485.ch3.

9. Leventhal H, Leventhal EA, Contrada RJ. Self-regulation, health, and behavior: A perceptual-cognitive approach. Psychol Health. 1998; 13(4): 717–733 10.1080/08870449808407425.

10. Moss-Morris R, Weinman J, Petrie K, Horne R, Cameron L, Buick D. The Revised Illness Perception Questionnaire (IPQ-R). Psychol Health. 2002; 17(1): 1–16 10.1080/08870440290001494.

11. Mora PA, McAndrew LM. Common-Sense Model of Self-regulation. In: Gellman MD, Turner JR, (eds.) Encyclopedia of behavioral medicine. New York: Springer 2013. p. 460–467. 10.1007/978-1-4419-1005-9_1220.

12. Ritchie J, Spencer L. Qualitative data analysis for applied policy research. In: Bryman A, Burgess B, (eds.) Analyzing qualitative data. London, UK: Routledge 1994. p. 173–194.

13. Segal DL, June A, Payne M, Coolidge FL, Yochim B. Development and initial validation of a self-report assessment tool for anxiety among older adults: The Geriatric Anxiety Scale. J Anxiety Disord. 2010; 24(7): 709–714 10.1016/j.janxdis.2010.05.002.

14. Yesavage JA, Sheikh JI. Geriatric Depression Scale (GDS): Recent evidence and development of a shorter version. Clin Gerontol. 1986; 5(1-2): 165–173 10.1300/J018v05n01_09.

15. Taqui AM, Itrat A, Qidwai W, Qadri Z. Depression in the elderly: Does family system play a role? A cross-sectional study. BMC Psychiatry. 2007; 7(1): 57 10.1186/1471-244X-7-57.

16. Prakash O, Gupta LN, Singh VB, Nagarajarao G. Applicability of 15-item Geriatric Depression Scale to detect depression in elderly medical outpatients. Asian J Psychiatr. 2009; 2(2): 63–65 10.1016/j.ajp.2009.04.005.

17. Lahiri A, Chakraborty A. Psychometric validation of Geriatric Depression Scale - Short Form among Bengali-speaking elderly from a rural area of West Bengal: Application of item response theory. Indian J Public Health. 2020; 64(2): 109–115 10.4103/ijph.IJPH_162_19.

18. Ritchie J, Lewis J. Qualitative research practice: A guide for social science students and researchers. London: Sage Publications; 2003.

19. Goldsmith L. Using Framework Analysis in applied qualitative research. The Qualitative Report. 2021; 26(6): 2061–2076 10.46743/2160-3715/2021.5011.

20. Gale NK, Heath G, Cameron E, Rashid S, Redwood S. Using the Framework Method for the analysis of qualitative data in multi-disciplinary health research. BMC Med Res Methodol. 2013; 13: 10.1186/1471-2288-13-117.

21. Nair P, Bhanu C, Frost R, Buszewicz M, Walters KR. A systematic review of older adults’ attitudes towards depression and its treatment. Gerontologist. 2020; 60(1): e93–e104 10.1093/geront/gnz048.

22. Corcoran J, Brown E, Davis M, Pineda M, Kadolph J, Bell H. Depression in older adults: A meta-synthesis. Journal of Gerontological Social Work. 2013; 56(6): 509–534 10.1080/01634372.2013.811144.

23. Kendell R, Jablensky A. Distinguishing between the validity and utility of psychiatric diagnoses. Am J Psychiatry. 2003; 160(1): 4–12 10.1176/appi.ajp.160.1.4.

24. Leventhal H, Phillips LA, Burns E. The Common-Sense Model of Self-Regulation (CSM): A dynamic framework for understanding illness self-management. J Behav Med. 2016; 39(6): 935–946 10.1007/s10865-016-9782-2.

25. Wiedemann K. Anxiety and anxiety disorders. In: Smelser NJ, Baltes PB, (eds.) International encyclopedia of the social & behavioral sciences. Elsevier 2001. p. 560–567. 10.1016/B0-08-043076-7/03760-8.

26. Rose G, Barker DJ. Epidemiology for the uninitiated. What is a case? Dichotomy or continuum? Br Med J. 1978; 2: 873–874 10.1136/bmj.2.6141.873.

27. Drzewiecki CM, Fox AS. Understanding the heterogeneity of anxiety using a translational neuroscience approach. *Cognitive, Affective*, & Behavioral Neuroscience. 2024; 24(2): 228–245 10.3758/s13415-024-01162-3.

28. Dowrick C. Beyond depression: A new approach to understanding and management. 2nd ed. Oxford: Oxford University Press; 2009.

29. Kinderman P, Setzu E, Lobban F, Salmon P. Illness beliefs in schizophrenia. Soc Sci Med. 2006; 63(7): 1900–1911 10.1016/j.socscimed.2006.04.022.

30. Oris L, Luyckx K, Rassart J, Goubert L, Goossens E, Apers S, et al. Illness identity in adults with a chronic illness. J Clin Psychol Med Settings. 2018; 25(4): 429–440 10.1007/s10880-018-9552-0.

31. Wierzbicka A. Human emotions: Universal or culture-specific? American Anthropologist. 1986; 88(3): 584–594 10.1525/aa.1986.88.3.02a00030.

32. Ashmore RD, Deaux K, McLaughlin-Volpe T. An organizing framework for collective Identity: Articulation and significance of multidimensionality. Psychol Bull. 2004; 130(1): 80–114 10.1037/0033-2909.130.1.80.

33. Thoits PA, Virshup LK. Me’s and we’s: Forms and functions of social identities. In: Richard DA, Jussim L, (eds.) Self and identity: Fundamental issues. Rutgers series on self and social identity, Vol. 1. New York: Oxford Academic; 1997. p. 106–134. 10.1093/oso/9780195098266.003.0005.

34. Linville PW. Self-complexity as a cognitive buffer against stress-related illness and depression. J Pers Soc Psychol. 1987; 52(4): 663–676 10.1037/0022-3514.52.4.663.

35. Matsumoto D, Juang L. Culture and psychology. 3rd ed. Australia Southbank: Wadsworth/Thomson Learning; 2004.

36. Adams G, Markus HR. Toward a conception of culture suitable for a social psychology of culture. In: Schaller M, Crandall CS, (eds.) The psychological foundations of culture. New Jersey: Lawrence Erlbaum Associates Publishers; 2004. p. 335–360.

## References

Gould CE, Segal DL, Yochim BP, Pachana NA, Byrne GJ, Beaudreau SA. Measuring anxiety in late life: A psychometric examination of the Geriatric Anxiety Inventory and Geriatric Anxiety Scale. J Anxiety Disord. 2014; 28(8): 804–811. doi:10.1016/j.janxdis.2014.08.001.

Yesavage JA & Sheikh JI. 9/Geriatric Depression Scale (GDS): Recent evidence and development of a shorter version. Clin Gerontol. 1986; 5(1–2), 165–173. 10.1300/J018v05n01_09

